# Methylphenidate stabilizes dynamic brain network organization during tasks probing attention and reward processing in stimulant-naïve children with ADHD

**DOI:** 10.1101/2025.01.27.25321175

**Authors:** Tehila Nugiel, Nicholas D. Fogleman, Margaret A. Sheridan, Jessica R. Cohen

## Abstract

Children with ADHD often exhibit fluctuations in attention and heightened reward sensitivity. Psychostimulants, such as methylphenidate (MPH), improve these behaviors in many, but not all, children with ADHD. Given the extent to which psychostimulants are prescribed for children, coupled with variable efficacy on an individual level, a better understanding of the mechanisms through which MPH changes brain function and behavior is necessary. MPH’s primary action is on catecholamines, including dopamine and norepinephrine. Catecholaminergic signaling can influence the tradeoff between flexibility and stability of brain function, which is one candidate mechanism through which MPH may alter brain function and behavior. Time-varying functional connectivity, which models how functional brain networks reconfigure on short timescales, can be used to examine brain flexibility versus stability, and is thus well-suited to test how MPH impacts brain function. Here, we scanned stimulant-naïve children with ADHD (8-12 years) on and off a single dose of MPH. In the MRI machine, participants completed two attention-demanding tasks: 1) a standard go/no-go task and 2) a rewarded go/no-go task. For both tasks, using a within-subjects design, we compared the degree to which brain organization changed throughout the course of the MRI scan, termed whole brain flexibility, on and off MPH. We found that whole brain flexibility decreased on MPH. Further, individuals with greater decreases in whole brain flexibility on MPH exhibited greater improvements in task performance. Together, these results provide novel insights into the neurobiological mechanisms underlying the effectiveness of MPH administration for children with ADHD.

## Introduction

Attention-deficit/hyperactivity disorder (ADHD) is the most common neurodevelopmental disorder, affecting approximately nine percent of children in the United States (Danielson et al., 2022). The primary symptoms of ADHD are inattention and hyperactive and impulsive behaviors. Foundational models of ADHD, such as the Dual Pathway Model, have proposed that ADHD symptoms arise from disruption to neurobehavioral circuits that orient attention through executive control and tune reward sensitivity through motivational control (Sonuga-Barke, 2002, 2003). Neuroimaging work in humans has confirmed that circuitry related to both executive control and motivational control is altered in individuals with ADHD (Costa Dias et al., 2015; Cubillo et al., 2012). Notably, these alterations are not limited to specific brain networks, but instead are widespread and include networks that support externally and internally focused attention, cognitive control, self-referential processing, and reward-related processing (da Silva et al., 2023; Mehta et al., 2019). Recent work has further identified that it is not only the organization of these networks, but how they dynamically change across time, that may be particularly relevant for understanding the etiology of ADHD (Cai et al., 2018; de Lacy & Calhoun, 2018; Duffy et al., 2021; Shappell et al., 2021; Yin et al., 2022).

Psychostimulants such as methylphenidate (MPH) are the most common first line intervention for ADHD (da Silva et al., 2023; Faraone, 2018). The primary mechanism of MPH is catecholaminergic modulation, through blocking dopamine transporters and elevating extracellular dopamine and norepinephrine (Faraone, 2018; Seeman & Madras, 2002). Preclinical studies of MPH action find that MPH-induced changes in extracellular dopamine and norepinephrine are pronounced in the prefrontal cortex (Berridge et al., 2006; Brozoski et al., 1979; Seamans & Yang, 2004) and at low doses lead to improved cognition and attention (Devilbiss & Berridge, 2008; Kodama et al., 2017). There is also evidence that MPH, through increased dopamine signaling, makes task-relevant stimuli more salient (Volkow et al., 2005), thus improving attention and goal-directed behavior (Volkow et al., 2004). Neuroimaging work in humans finds that acute MPH administration leads to widespread changes in functional connectivity in individuals with ADHD (da Silva et al., 2023; Silk et al., 2017; Yoo et al., 2018) and without ADHD (Farr et al., 2014; Mueller et al., 2014; Rosenberg et al., 2016; Siegel et al., 2023; Sripada et al., 2014); these changes have been linked to subsequent changes in behavior (Rosenberg et al., 2016). Despite growing evidence that MPH impacts functional connectivity averaged over several minutes, the impacts are heterogeneous across studies (Pereira-Sanchez et al., 2021). This may be because MPH targets the dynamics of functional connectivity, specifically how frequently functional connectivity patterns change across time.

A dynamic system, such as the brain, requires a balance between flexibility and stability (Liljenström, 2003; Safron et al., 2022). This balance supports different task demands, such as switching between tasks (flexibility) or sustaining attention to ignore distractions (stability; Armbruster et al., 2012). In terms of functional connectivity, this trade-off can be quantified by assessing the balance between maintaining a persistent functional connectivity pattern for a prolonged period of time (i.e., stability) and reconfiguration of this pattern at short timescales (i.e., flexibility). Dynamic functional connectivity methods can be used to study flexibility in functional connectivity patterns by modeling changes in functional connectivity on the order of seconds. In children with ADHD, dynamic functional connectivity during the resting state has been found to be decreased compared to children without ADHD (Yin et al., 2022), and to relate to errors on a go/no-go continuous performance task that probes sustained attention (Duffy et al., 2021). Also in children with ADHD, increased resting state dynamic functional connectivity has been found to relate to trait-level inattention symptoms (Cai et al., 2018). Finally, MPH administration in children with ADHD has been found to increase variability in dynamic functional connectivity patterns to be more similar to that of children without ADHD (Mizuno et al., 2022).

Dopamine signaling in the prefrontal cortex can shape brain flexibility (Cools, 2019) through a balance of dopamine D1 and D2 receptor signaling (Durstewitz & Seamans, 2008). In rats, MPH has been shown to change this balance in the prefrontal cortex by modulating D1 receptors and improving cognitive function (Arnsten & Dudley, 2005; Durstewitz & Seamans, 2008). However, it remains unknown how this change in the prefrontal cortex confers changes to whole brain functional network organization. Neuroimaging work in humans has observed that experimental dopamine depletion increases signal variability during the resting state distributed across functional brain networks (Shafiei et al., 2019), while catecholamine reuptake inhibitors have been shown to increase dynamic functional connectivity during working memory in healthy adults (Shine et al., 2018). Further, D2 receptor agonists have been shown to change orbitofrontal cortex functional connectivity with the rest of the brain (Kahnt & Tobler, 2017). These studies suggest that MPH may change whole brain function and dynamics via catecholaminergic modulation in the prefrontal cortex, however the direction and nature of this effect in children with ADHD needs clarification.

Here we propose to use dynamic functional connectivity methods to test the role of MPH in modulating brain flexibility as a potential mechanism through which MPH changes attention and reward processing in children with ADHD. We used an attention-demanding go/no-go paradigm and a go/no-go paradigm with performance-based rewards to examine how MPH changes dynamic functional connectivity on the order of seconds in stimulant-naïve children with ADHD. By testing how MPH-induced changes in dynamic functional connectivity and flexibility relate to both attention and reward processing behaviors, we can assess how MPH administration impacts multiple cognitive contexts that are typically disrupted in children with ADHD.

## Methods

Participants and eligibility: Study participants were a subsample from a larger study investigating how MPH changes brain network organization. The sample included in the current study were 36 children with ADHD, 8-12 years (mean age = 9.7 years, SD = 1.18, 17 females). All participants were psychostimulant medication-naïve at the time of the study. Participants’ race was White (30), Asian (2), Black (1), and Mulitracial (3). General exclusion criteria for the sample included the following: 1) full scale intelligence quotient (FSIQ) less than 85 as determined by the Wechsler Intelligence Scale for Children, Fifth Edition (Wechsler, 2014), and 2) Word Reading subtest score less than 85 as determined by the Wechsler Individual Achievement Test, Third Edition (Wechsler, 1992). Children were also excluded from the study if any of the following criteria were met: 1) diagnosis of intellectual disability, developmental speech/language disorder, reading disability, autism spectrum disorder, or a pervasive developmental disorder, 2) visual or hearing impairment, 3) neurologic disorder (e.g., epilepsy, cerebral palsy, traumatic brain injury, Tourette syndrome), or 4) medical contraindication to MRI (e.g., implanted electrical devices, dental braces). Children were included in the study sample if they met full diagnostic criteria for ADHD using the Diagnostic Interview Schedule for Children Version IV (DISC-IV; Shaffer et al., 2000) or if they reached a subthreshold diagnosis of ADHD with impairment on the DISC-IV and full criteria on the Conners 3rd Edition Parent or Teacher Rating Scales (Conners, 2008).

All participants underwent testing twice, once on MPH and once on placebo (see *Study procedure* for details). Since our primary interest in the current study was examining the effects of MPH, only individuals who had data for both MPH and placebo sessions for at least one task were included in our analyzed sample. After excluding fMRI runs for quality control (see *fMRI processing*), 31 participants (mean age = 9.72 years, SD = 1.14, 14 females, White (26), Asian (2), Black (1), and Mulitracial (2)) were included in analyses. Of the 31 participants, 28 were included in the standard go/no-go task analyses and 29 were included in the rewarded go/no-go task analyses.

### Study procedure

All study procedures were reviewed and approved by the Institutional Review Board at The University of North Carolina at Chapel Hill. Parental informed consent and child assent was obtained for all participants. Approximately one week prior to the MRI scans, participants underwent a mock scanning procedure in which they were introduced to the scanner setting and trained to remain still with a combination of real-time motion tracking and incentives. Participants also practiced the go/no-go tasks and underwent other assessments that will not be discussed here. Following this initial visit, participants were scanned twice, once on MPH and once on placebo, approximately one week apart in a double-blind single-dose procedure in which MPH/placebo order was randomized. MPH and placebo were administered one hour before scanning in identical form. An MPH dose of 0.3 mg/kg was administered; this is a safe initial dose that has been used in prior research with medication-naïve children with ADHD (Faraone, 2018; Rubia et al., 2009) and has been shown to change brain function and improve task performance in children with ADHD (Rubia et al., 2009, 2011).

### fMRI tasks

We tested the effects of MPH on attention and reward processing using a standard go/no-go task and a rewarded go/no-go task.

#### Standard go/no-go task

Participants viewed one of eight sports balls presented on an MRI-compatible projector one at a time (**Figure 1a**). Two of the eight sports balls were randomly selected as ‘no-go’ stimuli for each participant. The remaining 6 sports balls were ‘go’ stimuli. Participants were told to press the index finger of their right hand when ‘go’ sports balls were on the screen (∼73% of trials) and to withhold from pressing when ‘no-go’ sports balls were on the screen (∼27% of trials). Each of two standard go/no-go runs consisted of 128 trials. Sports balls were displayed for 600ms with a jittered interstimulus interval ranging from 1.25 to 3.25s.

**Figure 1.**
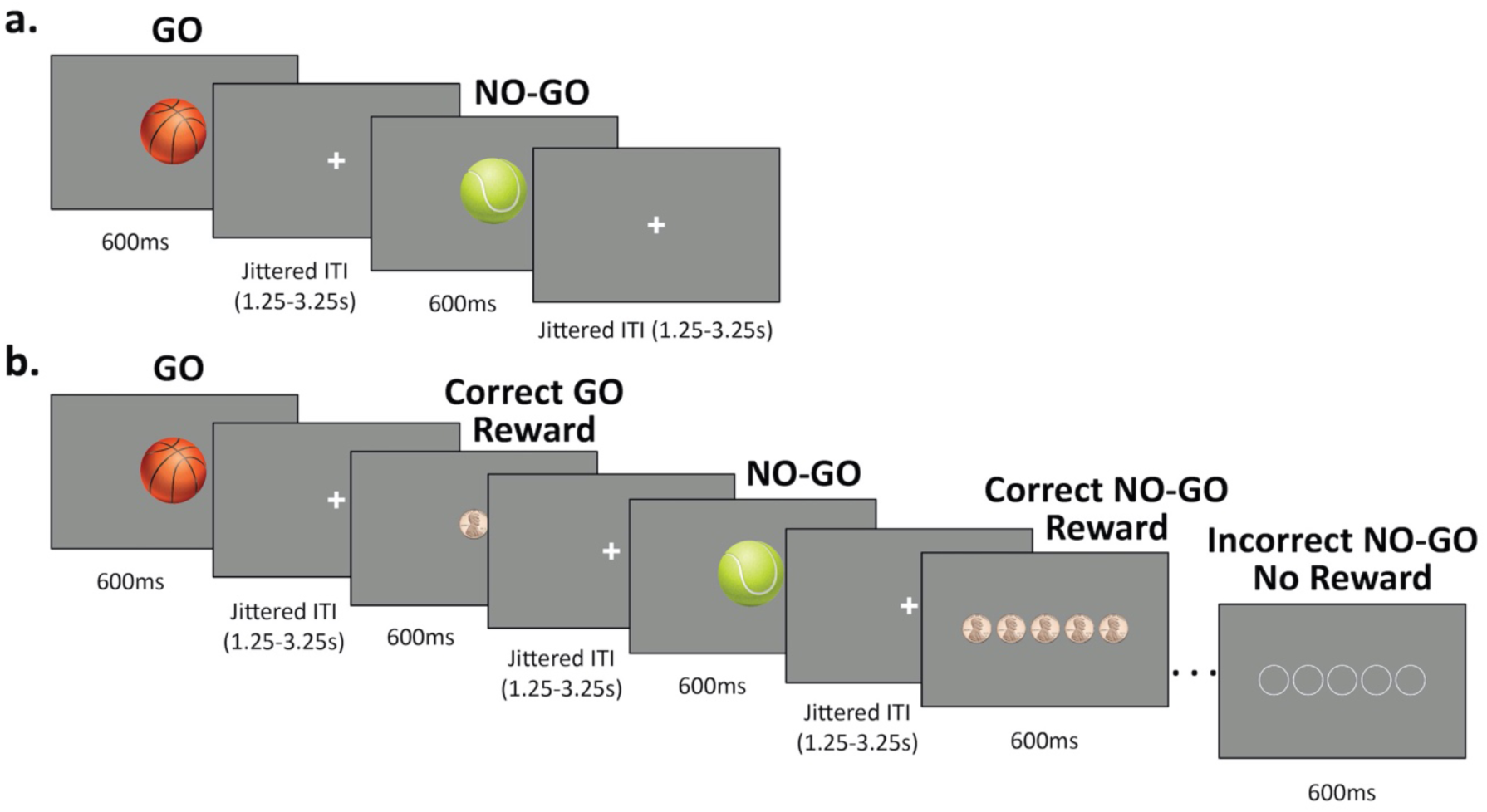
fMRI Task stimuli. **a.** Standard go/no-go task: Sports balls were presented one at a time. In this example, basketballs were ‘go’ stimuli and the correct response was a button press. Tennis balls were ‘no-go’ stimuli, and the participant was instructed to withhold pressing the button. **b.** Rewarded go/no-go task: Each sports ball stimulus was followed by the reward that was earned (pennies shown) or not earned (empty circles shown). The potential reward for each go trial was one penny. The potential reward for each no-go trial was five pennies.

#### Rewarded go/no-go task

The rewarded version of the go/no-task was identical to the standard version of the go/no-go task, with the addition of reward feedback presented after each trial based on performance (**Figure 1b**). Participants were told that correct, fast responses on go trials (≤650 ms) and correct non-responses on no-go trials would be rewarded. Each of four rewarded go/no-go runs consisted of 64 trials. Stimulus presentation and timing were identical to those of the standard go/no-go task, with the addition of feedback displayed after each response. The interval between the stimulus and the reward feedback matched the jittered interstimulus interval between trials (range 1.25-3.25s). Reward feedback following the jitter was presented for 600ms. Participants were presented with an image of one penny for correct responses on go trials and five pennies for correctly withheld responses on no-go trials. The same number of empty circles appeared on trials that were not rewarded. Money was accumulated throughout the scan and given to participants at the end of the session.

To assess fluctuations in attention, we calculated tau as a measure of response time variability. Tau was calculated using the *retimes* package in R (Massidda, 2013) as the exponential component of the response time distribution fitted with an exponential-Gaussian (ex-Gaussian) curve (Balota & Yap, 2011). To assess task accuracy, we calculated d’, which was calculated as the difference between distribution functions of hit rate (i.e., proportion of correct go trials) and false alarm rate (i.e., proportion of commission errors) (Stanislaw & Todorov, 1999).

### MRI collection protocol

MRI data were collected on a 3-tesla Siemens MAGNETOM Prisma-fit whole-body MRI machine with a 32-channel head coil at the University of North Carolina at Chapel Hill Biomedical Research Imaging Center. A high-resolution anatomical T1-weighted MPRAGE scan was collected (TR = 2400 ms, TE = 2.22 ms, FA = 8°, field of view 256mm x 256mm, 208 slices, in-plane voxel size 0.8 mm). Whole brain functional data were collected with a gradient-echo T2*-weighted EPI sequence (39 slices, TR = 2000 ms, TE = 25 ms, flip angle = 77°, echo spacing = 0.54 ms, field of view 230 × 230 mm, voxel dimensions 2.9 mm × 2.9 mm × 3 mm). Two runs of approximately 6.5 minutes each (195 TRs) were collected of the standard go/no-go task, and 4 runs of approximately 6.16 minutes each (185 TRs) were collected of the rewarded go/no-go task. Due to a randomly sampled jitter, task length varied slightly across participants.

### fMRI processing

fMRI data preprocessing was performed using fMRIPrep 21.0.2 (Esteban et al., 2019), with steps including EPI to T1w coregistration, susceptibility artifact correction, normalization to MNI space, and estimation of motion parameters. Preprocessed runs were visually quality assessed and runs with significant artifacts or runs that included ROIs from the final ROI set with < 50% coverage of brain data were excluded from further analyses. ICA AROMA (Pruim et al., 2015) was used during preprocessing to identify spatial-temporal noise components that were later removed (see below). Preprocessing details can be found in the *Supplemental Materials*. In addition to ICA AROMA denoising, functional runs with a mean framewise displacement > 0.5 mm were excluded from analyses (19 runs from 10 participants). After preprocessing, the following steps took place. First, confound signals from cerebrospinal fluid, white matter, and global signal, as well as their temporal, quadratic, and quadratic temporal derivatives calculated from fMRIPrep, were generated. Then, the noise components identified with AROMA were non-aggressively regressed out of both the data and the confounds. Data and confounds were then temporally filtered with a high pass filter (0.08 mHz). Next, the AROMA-regressed and filtered confound signals were regressed from the data. Lastly, a brain mask created with fMRIPrep’s MCFLIRT step was applied to the data to remove non-brain tissue from the image.

After processing, a whole brain functional atlas composed of 300 5 mm radius spherical regions of interest (ROIs) (Seitzman et al., 2020) was applied to the data and average timeseries of activity for each ROI were extracted. ROIs that did not belong to a specific network were removed from further analysis (N = 12), as were additional ROIs that fell outside of the brain masks for more than 10 runs (N = 21), leaving a timeseries of 267 ROIs. For a table totaling how many runs each participant contributed to the final dataset, see the *Supplemental Materials*.

### Dynamic conditional correlation

To estimate time-varying functional connectivity of the 267 ROIs we used dynamic conditional correlation (DCC; Choe et al., 2017; Lindquist et al., 2014). DCC is a multivariate volatility model based on a generalized autoregressive heteroscedastic approach (Lindquist et al., 2014). A conditional covariance is computed between all pairs of ROIs at a given timepoint, resulting in one correlation matrix per timepoint (every two seconds). The covariances are computed based on a weighted combination of past values with a window length determined using a quasi-maximum likelihood approach. DCC has been found to be more reliable in capturing time-varying covariance structure than other sliding window approaches (Choe et al., 2017; Lindquist et al., 2014). A 267 x 267 correlation matrix for each timepoint was constructed with the first and last 5 volumes discarded to allow the model to stabilize. Each correlation matrix was Fisher z-transformed. This resulted in ∼178 267 x 267 correlation matrices per participant per run of the standard go/no-go task, and ∼170 per participant per run of the rewarded go/no-go task.

### Multilayer networks and whole brain flexibility

We used multilayer networks (Bassett et al., 2013; Mucha et al., 2010) to assess dynamic brain organization. Each matrix in the DCC series of 267 x 267 matrices formed a layer for a multilayer network (Mucha et al., 2010). Multilayer modularity (Bassett et al., 2013; Betzel et al., 2019; Mucha et al., 2010) was used to model functional organization in each layer as well as reconfiguration from layer to layer. Modularity characterizes the community structure of a network wherein communities of nodes have stronger within-community than between-community connections. Community structure is defined by maximizing modularity at any given layer, thus community assignment of a given node can change across layers. Multilayer modularity requires specification of two free parameters: ***γ***, which modulates interlayer connections and ***Ω***, which modulates intralayer connections (Bassett et al., 2011; Mucha et al., 2010). We set the parameters at ***γ*** = 1.5 and ***Ω*** = 1, which gave us 10.96 communities on average in each layer (range 6-16), which closely recapitulated the number of included communities in the functional brain atlas we used (N=12).

Our primary metric of interest was flexibility (Telesford et al., 2016), which is a measure of how often a node changes community membership across layers. Flexibility is a fractional metric ranging from 0-1, calculated as the number of times a node switches community assignment across layers normalized by the number of possible changes (Bassett et al., 2011; Telesford et al., 2016). We quantified whole brain flexibility by averaging flexibility across all 267 nodes, with higher flexibility indicating more changes in brain organization (i.e., community membership) across time. Whole brain flexibility was not related to framewise displacement (r = 0.02, p = 0.72), indicating that our processing successfully removed the impact of head motion on functional connectivity estimates (Satterthwaite et al., 2012; Siegel et al., 2014).

### Analytic models

We used Pearson correlations to test the relationship between our two task performance metrics, tau and d’. We used mixed effects models with a random effect of subject and fixed effect of condition (placebo/MPH) to test differences in attention (tau), accuracy (d’), and whole brain flexibility on as compared to off MPH. We ran these models separately for the standard and rewarded go/no-go tasks. To index MPH-related change in brain and behavior we calculated the difference between the two conditions (MPH - placebo) for change in whole brain flexibility, change in tau, and change in d’ during each task. Changes in whole brain flexibility were then correlated with changes in task performance to test for the relationship between changes in whole brain flexibility and changes in behavior on MPH. All *p*-values were corrected using the false discovery rate (FDR) method (Benjamini & Hochberg, 1995) for the number of tests run in the corresponding analysis.

## Results

### MPH improved task performance

On placebo, tau and d’ were negatively correlated with each other during both the standard go/no-go task (*r* (26) = -0.67, corrected-p < .0001) and the rewarded go/no-go task (*r* (27) = - 0.77, corrected-p < .0001).

On MPH, tau and d’ were negatively correlated with each other during the standard go/no-go task (*r* 26) = -0.48, corrected-p =.02), but not during the rewarded go/no-go task (*r* (27) = -0.3, corrected-p =.11). *P*-values were FDR corrected for two tests.

During both go/no-go tasks, performance improved on MPH. During the standard go/no-go task, tau was higher on placebo as compared to MPH (*b* = 0.023, corrected-*p* = 0.035; mean placebo = 0.13; mean MPH = 0.12) and d’ in was lower on placebo as compared to MPH (*b* = -0.28, corrected-*p* = 0.027; mean placebo = 1.86; mean MPH = 2.09). Similarly, during the rewarded go/no-go task, tau was higher (*b* = 0.033, corrected-*p* = 0.002; mean placebo = .13; mean MPH = .10) and d’ was lower (*b* = -0.52, corrected-*p* = 0.001; mean placebo = 2.03; mean MPH = 2.53) on placebo as compared to MPH. Change in behavior on MPH, particularly for d’, was numerically larger during the rewarded compared to the standard go/no-go task, although there was not a significant interaction between change in behavior and task condition (uncorrected-*p* = 0.49 for tau, and uncorrected-*p* = 0.20 for d’).

### MPH decreased whole brain flexibility

Whole brain flexibility was lower on MPH than on placebo during the standard go/no-go task (Figure 2a; *b*= 0.0045, corrected-*p* = 0.01; mean placebo = .048, mean MPH = .043) and during he rewarded go/no-go task (Figure 2b; *b* = 0.005, corrected-*p* = 0.0009, mean placebo = .048, mean MPH = .042). While the effect was numerically stronger during the rewarded go/no-go task, there was not a significant interaction between change in whole brain flexibility and task condition (uncorrected-*p* = 0.85).

**Figure 2.**
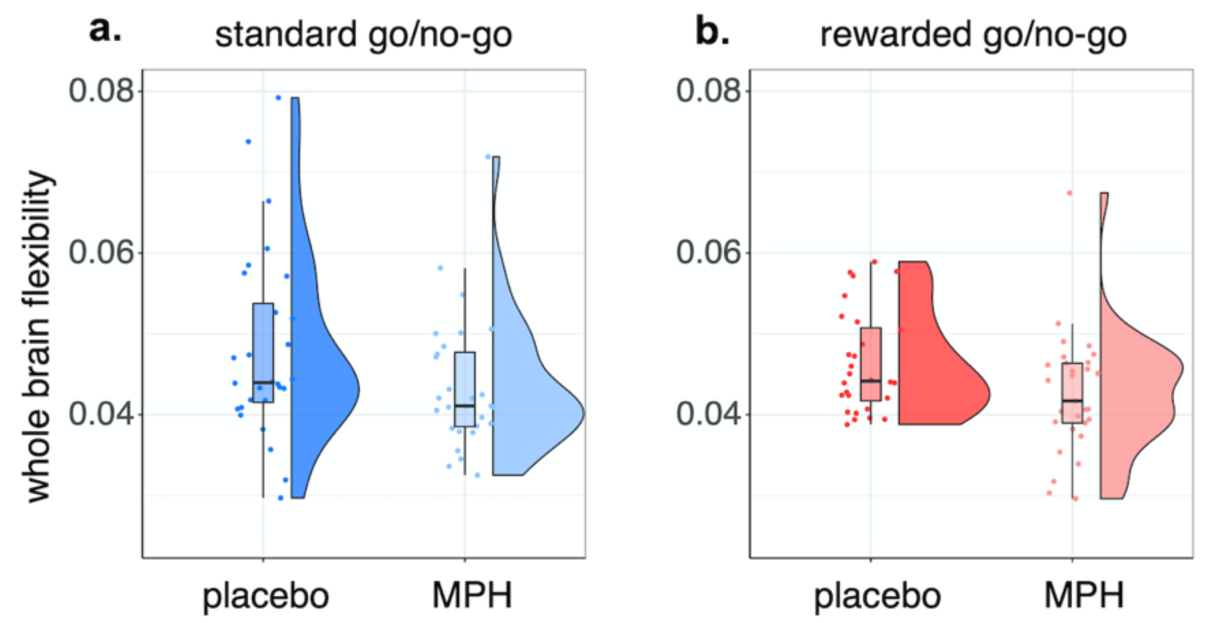
Whole brain flexibility decreased on MPH a) during the standard go/no-go task (*b*= 0.0045, corrected-p = 0.01) and b) during the rewarded go/no-go task (*b*= 0.005, corrected-p = 0.0009).

### Changes in task performance were related to changes in whole brain flexibility

Finally, we determined the relationship between the change in whole brain flexibility and changes in both tau and d’. Change in whole brain flexibility was positively correlated with change in tau during both go/no-go tasks (standard go/no-go: *r(*26) = 0.43, corrected-*p* = .032; Figure 3a; rewarded go/no-go: *r(*27) = 0.43, corrected-*p* = .032; Figure 3b). Specifically, larger decreases in whole brain flexibility were related to larger decreases in tau. Change in whole brain flexibility was negatively correlated with change in d’ during the rewarded go/no-go task (*r(*27) = -0.47, corrected-*p* = .032, Figure 3c). Specifically, larger decreases in whole brain flexibility were related to larger increases in d’. This relationship was not significant in the standard go/no-go task (*r(*26) = -0.03, *p* = 0.87) though the difference between the correlations did not reach significance (*p* = .09).

**Figure 3.**
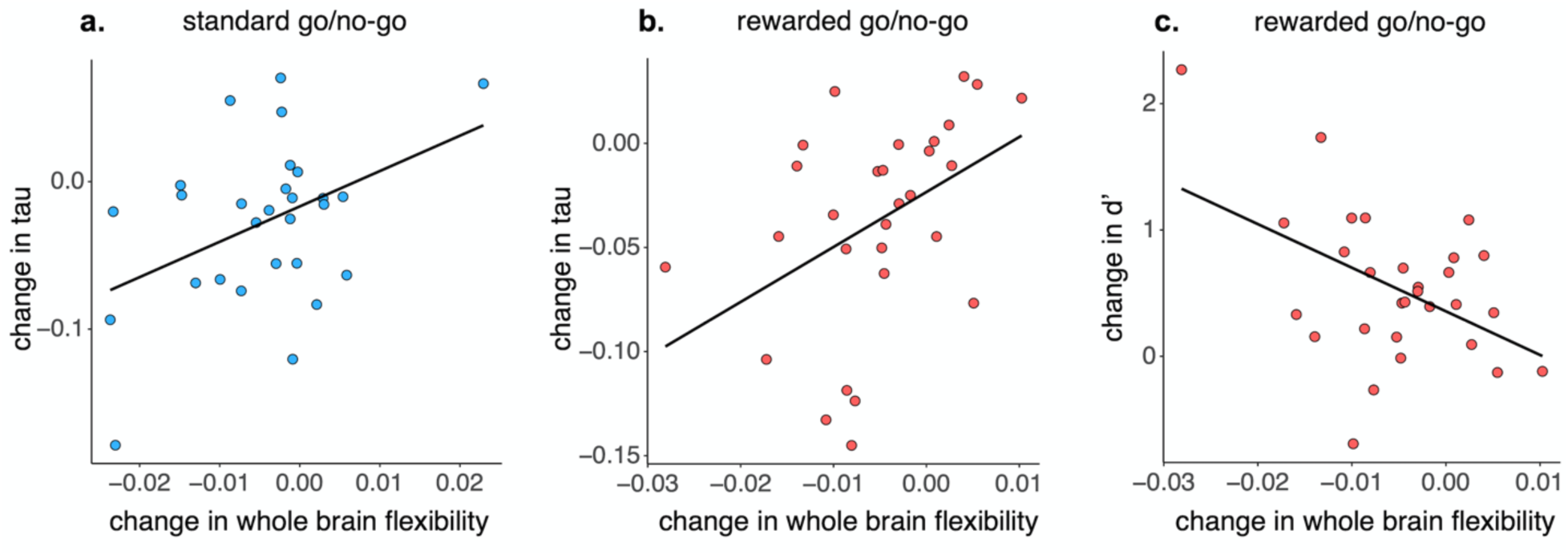
Change in whole brain flexibility on MPH was related to a) change in tau during the standard go/no-go task (*r*(26) = 0.43, corrected-p = 0.032), b) change in tau during the rewarded go/no-go task (*r*(27) = 0.43, corrected-p = 0.032), and c) change in d’ during the rewarded go/no-go task (*r*(27) = -0.47, corrected-p = 0.032). Change for all measures was calculated as MPH-placebo.

## Discussion

We examined how a single dose of MPH changed whole brain flexibility in a sample of stimulant-naïve children with ADHD. This study builds on work asserting that catecholaminergic signaling in the prefrontal cortex plays a role in ADHD by disrupting the balance between neural flexibility and stability (Arnsten, 2009), and that MPH improves cognitive function through increasing prefrontal catecholaminergic signaling (Arnsten & Dudley, 2005). We extend these findings to examine signaling across the whole brain by assessing network-level flexibility in children with ADHD as a means to assess how MPH changes the global balance between flexibility and stability. We observed that whole brain flexibility decreased on MPH during both a standard and a rewarded go/no-go task. Importantly, this decrease was related to a decrease in tau, a measure of response time variability, consistently across the standard and rewarded go/no-go tasks, and to an increase in d’ during the rewarded go/no-go task only.

### Methylphenidate targets the balance between flexibility and stability

A dual-state theory of dopamine function in the prefrontal cortex posits two states dominated by D1 and D2 receptors that impact prefrontal network dynamics (Durstewitz & Seamans, 2008). Imbalance of these states has been posited to lead to cognitive disruptions. This forms a neurobiological hypothesis of ADHD (Arnsten, 2009), wherein disrupted D1 receptor function increases ‘noise’ and weakens stability in prefrontal cortex dynamics. Therapeutic doses of MPH have been shown to change dopamine modulated neuronal firing in the prefrontal cortex (Berridge et al., 2006; Kuczenski & Segal, 1997), partially through D1 receptor stimulation (Arnsten & Dudley, 2005), and to improve cognitive function (Arnsten & Dudley, 2005). Beyond prefrontal functioning, it has been asserted that the balance between flexibility and stability is important for the efficient function of large scale brain networks distributed across the entire brain (Safron et al., 2022). Building from this assertion, our work extends the prefrontal-focused neurobiological hypotheses of MPH in ADHD by measuring variability in blood oxygen level dependent signal throughout the entire brain. We hypothesize that the change in prefrontal dynamics has cascading effects via functional connections distributed throughout the brain. Consistent with our hypothesis, we found that MPH decreased whole brain flexibility (i.e., reduced variability and increased stability), corroborating theories stating that MPH ameliorates a tendency toward network instability in ADHD.

Previous work has reported that in youth with ADHD with a history of psychostimulant use, variability in dynamic functional connectivity estimates is increased (Yin et al., 2022), and acute MPH administration increases that variability further (Mizuno et al., 2022). While the direction of these findings may seem to contrast our results, there are several important distinctions that make our results complementary to this literature. First, our work administers an acute dose of MPH to children who are psychostimulant naïve, while previous work has focused on youth with a history of psychostimulant use and thus speaks to chronic effects of MPH. Further, our work examines whole brain flexibility during attention demanding tasks, while previous work examined brain dynamics at rest. Our findings align with our recent work in a different sample reporting that children with ADHD showed increased dynamic functional connectivity variability compared to youth without ADHD during go/no-go tasks (Michael et al., 2024). Together, this body of work highlights that MPH effects can vary by cognitive context (rest vs. task) and child context (acute vs. chronic users of MPH), clarifying important nuances related to MPH use in children with ADHD.

In addition to its primary action on dopamine signaling, MPH also increases extracellular norepinephrine through blocking norepinephrine transporters (Hannestad et al., 2010). This change in norepinephrine signaling could be another pathway through which MPH changes large-scale brain dynamics, through the locus coeruleus, the primary source of cortical norepinephrine (Bouret & Sara, 2005). Similar to dopamine signaling in the prefrontal cortex discussed above, imbalance of a1 and a2 norepinephrine receptors in the prefrontal cortex can also impair cognition (Berridge & Spencer, 2016). The relationship between prefrontal norepinephrine receptors and flexibility is complex, as a1 receptors have been found to promote both focused and flexible attention (Berridge et al., 2011). Taken together, the effects on norepinephrine signaling are important to consider alongside dopamine signaling as mechanisms of stimulant-driven change on brain dynamics and behavior. The lack of specificity of MPH and our fMRI approach does not allow us to tease apart the effects of dopamine versus norepinephrine signaling in this study, but future work using specific transporter blockers or neurochemical imaging can make these important distinctions.

### Individual differences in the degree of change in brain and behavior after MPH administration

Psychostimulants, such as MPH, work well in many children with ADHD, but ∼20-30% do not do not see notable improvements in attention and behavior (Cortese, 2020; Pagnier, 2023). Consistent with these statistics, while MPH decreased whole brain flexibility on a group level in the current sample, we also observed marked individual differences in the degree, and even the direction, of change in whole brain flexibility on MPH. These individual differences in how MPH altered whole brain flexibility were related to individual differences in change in task performance. Tau quantifies particularly slow responses in an ex-Gaussian distribution (Balota & Yap, 2011), which is thought to reflect fluctuations in attention (Karalunas et al., 2014; Leth-Steensen et al., 2000), and is higher in individuals with ADHD on a group level (Karalunas et al., 2014; Lin et al., 2015). We observed that decreases in whole brain flexibility on MPH were correlated with decreases in tau, or decreased response time variability, on MPH. In other words, MPH drove both the brain and behavior into a more stable state. In addition to lower tau on MPH, reductions in whole brain flexibility were also related to increases in task accuracy, indexed by d’, during the rewarded go/no-go task. Both MPH and the receipt of rewards have been shown to improve cognitive performance in children with ADHD (Rosch et al., 2016) through their modulation of dopaminergic signaling (Arnsten & Rubia, 2012). Together, rewards and MPH have an additive augmenting effect on cognitive performance (Rosch et al., 2016). Taken together, the current work indicates that MPH may change attention and task accuracy by modulating whole brain flexibility.

As stated above, we observed that MPH changed whole brain network dynamics in a way that was adaptive for many participants in this study, but perhaps maladaptive for a smaller subset. This provides a possible explanation for why stimulants do not reduce symptoms in some individuals with ADHD. This suggests that individuals in our study initially fell along different points of the inverted-U that has been used to describe the relationship between dopamine signaling and the balance of flexibility/stability (Cools & D’Esposito, 2011). This is evidenced by our previous work examining T2* relaxation rate in the current study’s sample as a proxy for dopamine availability, which found marked variability in dopamine availability (Cascone et al., 2023). Thus, while MPH shifted the brain toward stability in many participants, it shifted the brain toward flexibility and poorer task performance in others. Differential neural and behavioral responses, including those that may be maladaptive, have important clinical implications. We consider our individual differences analyses preliminary due to sample size, and look forward to future studies in larger samples that can replicate and expand upon these findings to better understand MPH action in children with ADHD.

### Impact and conclusion

MPH is a widely prescribed psychostimulant for children with ADHD (Cortese, 2020), with increasing use in recent years (Chai et al., 2024). Yet it is not uniformly effective (Cortese, 2020; Pagnier, 2023), necessitating a better understanding of how MPH changes brain function and behavior, and in which individuals it is most likely to be effective. Here, we conducted an MPH challenge in a psychostimulant-naïve sample of children with ADHD, coupled with estimations of dynamic functional connectivity and performance on tasks probing attention and reward processing, to test how MPH changes whole brain flexibility and behavior. We observed that, on a group level, MPH decreased whole brain flexibility and improved attention during both go/no-go and rewarded go/no-go tasks. MPH additionally improved overall task accuracy, but only during the rewarded go/no-go task. Reduced MPH-induced flexibility was coupled with better task performance. Conversely, greater MPH-induced flexibility, observed in a subset of individuals, was coupled with poorer performance. Together, this research provides evidence that a mechanism through which MPH improves cognition and reduces primary ADHD symptoms may be through stabilizing whole brain network dynamics. Though we did not directly assess dopamine or norepinephrine availability, our findings extend previous prefrontal-focused theories of MPH action to large-scale brain dynamics. This work provides novel experimentally-derived insight into how MPH changes large-scale brain function and cognition and improves our understanding of MPH as a first-line treatment for children with ADHD.

## Data Availability

Data and code used in this manuscript is available upon request to the senior author (JRC).

## Acknowledgements -

Many thanks to Peter Mucha for developing the tools used for this work and for his support and time in helping us use them. We thank Kelly Eom and Cheyenne Bricken for their effort in data collection and project administration. We thank Laura Polittle and Diana Cejas for their support in conducting the study. Lastly, we thank the participants and their families for their time and effort.

