## Supplemental Materials for "Methylphenidate stabilizes dynamic brain network organization during tasks probing attention and reward processing in stimulant-naïve children with ADHD"

**Table 1. The number of subjects contributing each number of runs to standard go/no-go analyses after runs were excluded based on quality control criteria.**

| **Number of subjects** | **Number of standard**  **go/no-go runs MPH** | **Number of standard**  **go/no-go runs placebo** |
| --- | --- | --- |
| 1 | 1 | 1 |
| 4 | 1 | 2 |
| 6 | 2 | 1 |
| 17 | 2 | 2 |

**Table 2. The number of subjects contributing each number of runs to rewarded go/no-go analyses after runs were excluded based on quality control criteria.**

| **Number of subjects** | **Number of rewarded go/no-go runs MPH** | **Number of rewarded go/no-go runs placebo** |
| --- | --- | --- |
| 1 | 1 | 4 |
| 1 | 2 | 2 |
| 1 | 2 | 3 |
| 1 | 3 | 1 |
| 2 | 3 | 3 |
| 3 | 3 | 4 |
| 2 | 4 | 1 |
| 1 | 4 | 2 |
| 4 | 4 | 3 |
| 13 | 4 | 4 |

**fMRI processing details:**

Results included in this manuscript come from preprocessing performed using *fMRIPrep* 21.0.2 (Esteban, Markiewicz, et al., 2018; Esteban, Blair, et al., 2018; RRID:SCR_016216), which is based on *Nipype* 1.6.1 (K. Gorgolewski et al., 2011; K. J. Gorgolewski et al., 2018; RRID:SCR_002502).

**Anatomical data preprocessing**

A total of 1 T1-weighted (T1w) images were found within the input BIDS dataset.The T1-weighted (T1w) image was corrected for intensity non-uniformity (INU) with N4BiasFieldCorrection (Tustison et al., 2010), distributed with ANTs 2.3.3 (Avants et al., 2008, RRID:SCR_004757), and used as T1w-reference throughout the workflow. The T1w-reference was then skull-stripped with a *Nipype* implementation of the antsBrainExtraction.sh workflow (from ANTs), using OASIS30ANTs as target template. Brain tissue segmentation of cerebrospinal fluid (CSF), white-matter (WM) and gray-matter (GM) was performed on the brain-extracted T1w using fast (FSL 6.0.5.1:57b01774, RRID:SCR_002823, Zhang, Brady, and Smith, 2001). Brain surfaces were reconstructed using recon-all (FreeSurfer 6.0.1, RRID:SCR_001847, Dale, Fischl, and Sereno, 1999), and the brain mask estimated previously was refined with a custom variation of the method to reconcile ANTs-derived and FreeSurfer-derived segmentations of the cortical gray-matter of Mindboggle (RRID:SCR_002438, Klein et al., 2017). Volume-based spatial normalization to two standard spaces (MNI152NLin2009cAsym, MNI152NLin6Asym) was performed through nonlinear registration with antsRegistration (ANTs 2.3.3), using brain-extracted versions of both T1w reference and the T1w template. The following templates were selected for spatial normalization: *ICBM 152 Nonlinear Asymmetrical template version 2009c* [Fonov et al., 2009, RRID:SCR_008796; TemplateFlow ID: MNI152NLin2009cAsym], *FSL’s MNI ICBM 152 non-linear 6th Generation Asymmetric Average Brain Stereotaxic Registration Model* [Evans et al., 2012, RRID:SCR_002823; TemplateFlow ID: MNI152NLin6Asym].

**Functional data preprocessing**

For each of the 12 BOLD runs found per subject (across all tasks and sessions), the following preprocessing was performed. First, a reference volume and its skull-stripped version were generated using a custom methodology of *fMRIPrep*. Head-motion parameters with respect to the BOLD reference (transformation matrices, and six corresponding rotation and translation parameters) were estimated before any spatiotemporal filtering using mcflirt (FSL 6.0.5.1:57b01774, Jenkinson et al., 2002). BOLD runs were slice-time corrected to 0.964s (0.5 of slice acquisition range 0s-1.93s) using 3dTshift from AFNI (Cox and Hyde, 1997, RRID:SCR_005927). The BOLD time-series (including slice-timing correction) were resampled onto their original, native space by applying the transforms to correct for head-motion. These resampled BOLD time-series will be referred to as *preprocessed BOLD in original space*, or just *preprocessed BOLD*. The BOLD reference was then co-registered to the T1w reference using bbregister (FreeSurfer), which implements boundary-based registration (Greve and Fischl, 2009). Co-registration was configured with six degrees of freedom. Several confounding time-series were calculated based on the *preprocessed BOLD*: framewise displacement (FD), and three region-wise global signals. FD was computed using two formulations following Power (absolute sum of relative motions, Power et al.,2014) and Jenkinson (relative root mean square displacement between affines, Jenkinson et al., 2002). FD was calculated for each functional run, both using implementation in *Nipype* (following the definitions by Power et al., 2014). The three global signals were extracted within the CSF, the WM, and the whole-brain masks. The head-motion estimates calculated in the correction step were also placed within the corresponding confounds file. The confound time series derived from head motion estimates and global signals were expanded with the inclusion of temporal derivatives and quadratic terms for each (Satterthwaite et al., 2013). The BOLD time-series were resampled into standard space, generating a *preprocessed BOLD run in MNI152NLin2009cAsym space*. First, a reference volume and its skull-stripped version were generated using a custom methodology of *fMRIPrep*. Automatic removal of motion artifacts using independent component analysis (ICA-AROMA, Pruim et al., 2015) was performed on the *preprocessed BOLD on MNI space* time-series after removal of non-steady state volumes and spatial smoothing with an isotropic, Gaussian kernel of 6mm FWHM (full-width half-maximum). Additionally, the “aggressive” noise-regressors were collected and placed in the corresponding confounds file. All resamplings can be performed with *a single interpolation step* by composing all the pertinent transformations (i.e. head-motion transform matrices, and co-registrations to anatomical and output spaces). Gridded (volumetric) resamplings were performed using antsApplyTransforms (ANTs), configured with Lanczos interpolation to minimize the smoothing effects of other kernels (Lanczos, 1964). Non-gridded (surface) resamplings were performed using mri_vol2surf (FreeSurfer).

Many internal operations of *fMRIPrep* use *Nilearn* 0.8.1 (Abraham et al., 2014, RRID:SCR_001362), mostly within the functional processing workflow. For more details of the pipeline, see [the section corresponding to workflows in *fMRIPrep*’s documentation](https://fmriprep.readthedocs.io/en/latest/workflows.html).
